# Long-read sequencing with targeted assembly of the opsin locus accurately evaluates genes in expressed positions

**DOI:** 10.64898/2026.03.17.26348636

**Authors:** Zachary B. Anderson, Trent Prall, Nikhita Damaraju, Sophie HR Storz, Joy Goffena, Angela L. Miller, Joseph Carroll, Maureen Neitz, Danny E. Miller

## Abstract

The human opsin gene cluster at Xq28 contains highly similar *OPN1LW* and *OPN1MW* genes essential for red-green color vision. Current molecular methods cannot accurately analyze this complex locus, limiting diagnosis of color vision deficiencies (CVD) and detection of carrier status. We performed Nanopore long-read sequencing of 206 individuals, comparing alignment-based analysis with targeted *de novo* assembly. Alignment-based methods performed poorly, whereas targeted assembly achieved 99% concordance for *OPN1LW* and 92% for *OPN1MW* copy numbers and resolved gene order in all XY individuals and 87% of XX individuals. This approach detected CVD in 3.2% of XY individuals and identified 8% of XX individuals as carriers, consistent with population estimates. Moreover, it molecularly explained the phenotypic severity in a family with Bornholm eye disease and clarified carrier status in an XX individual suspected of carrying two CVD haplotypes. Our approach provides a comprehensive, reference-free method for accurate analysis of expressed opsin genes and reliable CVD carrier detection.

## INTRODUCTION

The opsin gene cluster located at Xq28 represents one of the most structurally complex regions of the human genome. This locus contains the *OPN1LW* and *OPN1MW* genes, which encode two opsin proteins responsible for human color vision—the long-wavelength (L) and middle-wavelength (M) cone opsins.^1,2^ These genes lie in a head-to-tail tandem array with a variable number of gene copies, typically consisting of an *OPN1LW* gene followed by one or more *OPN1MW* genes **(Figure 1A)**.^3^ The short-wavelength-sensitive (S) opsin gene (*OPN1SW*) is located within a nonrepetitive region of chromosome 7. Together, these three genes underlie normal trichromatic color vision.

**Figure 1.**
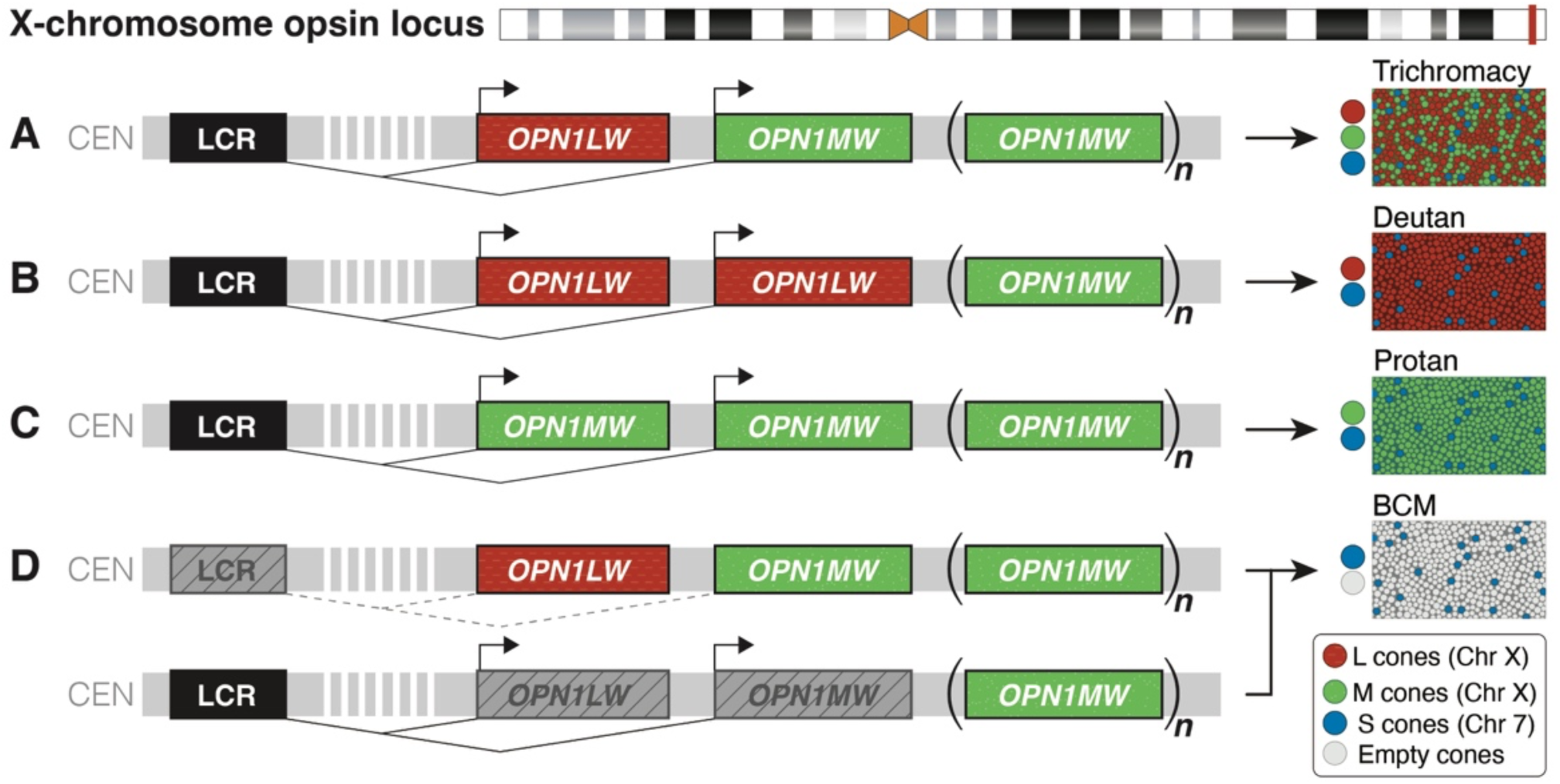
Structure of the opsin locus and impact of variation within the region. **(A)** The X-chromosome opsin locus is composed of a locus control region (LCR) that regulates expression of downstream opsin genes, typically beginning with *OPN1LW* (“L gene”, red cones) followed by one or more *OPN1MW* genes (“M gene”, green cones). Only the first two genes in the array are expressed, thus subsequent genes, if they are present, do not affect color vision. *OPN1LW* and *OPN1MW* are more than 98% identical at the coding sequence level. *OPN1SW* (“S gene”, not shown) resides on chromosome 7 and produces blue cones. Two common forms of color vision deficiency are (B) deutans, who have L genes in the two expressed positions, and (C) protans, who have M genes in the two expressed positions. (D) Loss of either the LCR or inactivating mutations in both of the first two genes in the array results in blue cone monochromacy (BCM), a slowly progressive cone dystrophy

Rearrangements affecting the order and number of opsin genes on the X chromosome are the primary cause of red-green color vision deficiencies (CVD) **(Figure 1B–D)**, which affect approximately 8% of XY individuals of European ancestry and 4–5% of XY individuals of African or Asian ancestry.^3^ Beyond their role in red-green color discrimination, variants in *OPN1LW* and *OPN1MW* are implicated in a spectrum of X-linked cone dysfunction disorders, including blue cone monochromacy, X-linked cone dystrophy, early-onset high myopia, and Bornholm eye disease.^4^

Transcription is regulated by a shared locus control region (LCR) located upstream of the gene array. In each cone photoreceptor cell, the LCR preferentially interacts with and activates only one of the first two genes closest to it. Consequently, normal trichromatic color vision requires that the first two genes in the array encode one L-class and one M-class pigment; alternative configurations can result in various forms of CVD. Because *OPN1LW* and *OPN1MW* transcription are mutually exclusive at the single-cell level, accurately determining the sequences of the first two genes in the array is essential for understanding color vision phenotypes.

Although the *OPN1LW* and *OPN1MW* genes share ≥98% sequence identity over ∼40 kilobases, the locus as a whole exhibits remarkable structural diversity among individuals. Accurate evaluation of sequence variants within these genes is critical. Spectral tuning differences between L- and M-class photopigments are primarily determined by amino acid polymorphisms at positions 277 and 285 in exon 5 **(Figure 2A,B)**. Whether an individual has normal trichromatic color vision or various forms of CVD depends on having one L-class and one M-class pigment encoded by the first two genes in the array. In addition, sequence polymorphisms within exon 3 at codons 153, 171, 174, 178, 180 influence the expression and splicing patterns of both genes, with specific polymorphisms leading to conditions like CVD, high myopia, or blue cone monochromacy **(Figure 2C)**.

**Figure 2.**
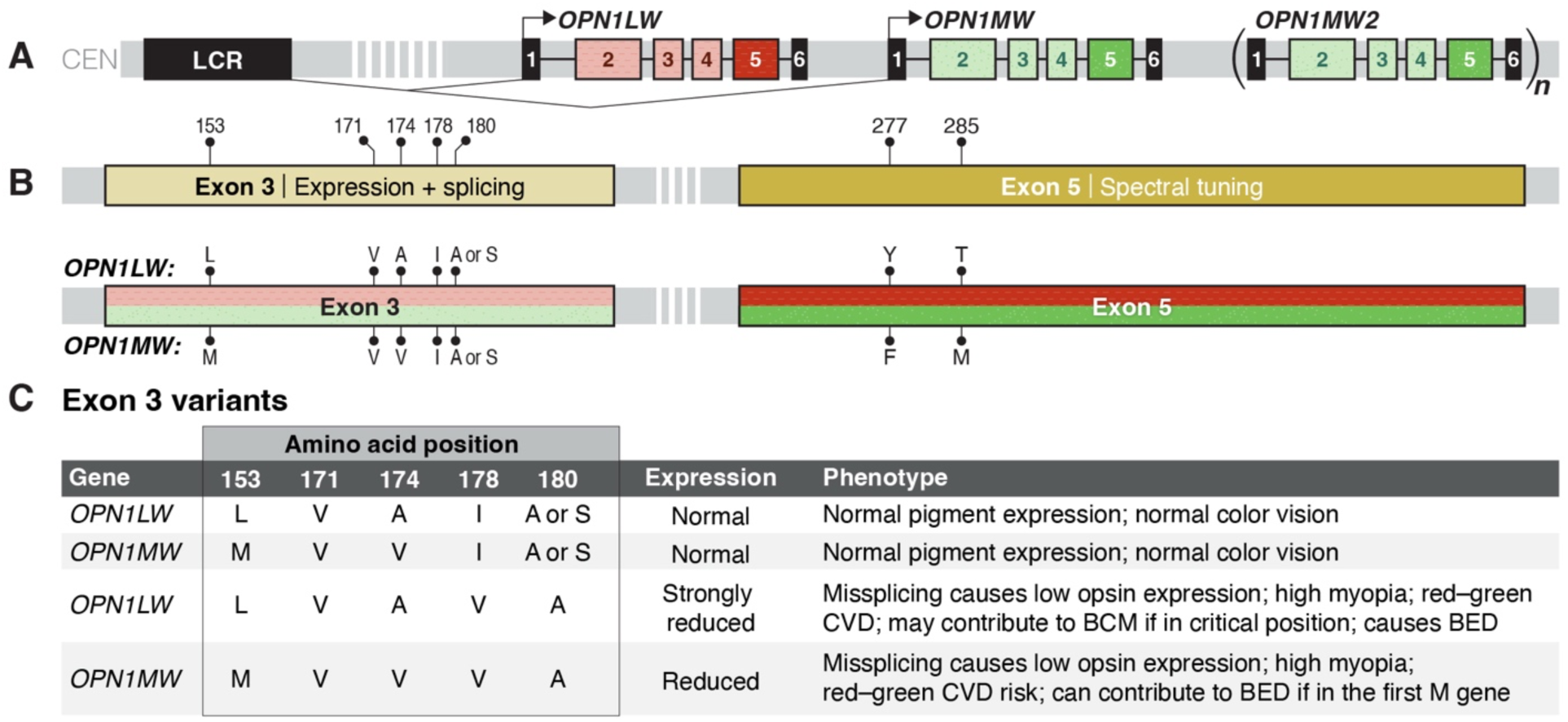
Sequence variation within exons 3 and 5 of *OPN1LW* and *OPN1MW*. **(A)** *OPN1LW* and *OPN1MW* are more than 98% identical at the coding sequence level, although intron 1 is ∼1,300 bp larger in *OPN1LW* than in *OPN1MW* in most individuals. **(B)** Polymorphisms at amino acid positions 277 and 285 in exon 5 are responsible for spectral tuning differences between L and M genes. Variation at amino acids 153, 171, 174, 178, and 180 in exon 3 affect expression and splicing, which may contribute to color vision deficiencies, myopia, and Bornholm eye disease. **(C)** Phenotypic effects of key exon 3 variants.

The complexity of this locus presents significant challenges for current molecular analysis methods. Short-read sequencing creates mapping ambiguities due to the high sequence similarity between L and M genes, while standard PCR-based methods are unreliable for complex arrays and ineffective in XX individuals. Quantitative and droplet digital PCR (ddPCR) can determine copy numbers but cannot resolve gene arrangement. Long-range PCR followed by long-read sequencing has shown promise but faces challenges with primer design in this repetitive region, and computational tools such as Paraphase may struggle with the extreme sequence similarity and produce ambiguous results in complex arrays or XX individuals.^5,6^ Collectively, these constraints have hindered comprehensive molecular evaluation of individuals with CVD, particularly for carrier detection in XX individuals.

Here, we present a comprehensive evaluation of the opsin locus using Oxford Nanopore (ONT) sequencing of 206 individuals from diverse populations sequenced by the 1000 Genomes Project Long-Read Sequencing Consortium (1KGP-LRSC) **(Supplemental Table 1)**.^7^ We compare alignment-based and assembly-based approaches and demonstrate that targeted *de novo* assembly achieves high concordance with orthogonal methods for determining *OPN1LW* and *OPN1MW* copy number, while also resolving the sequences of the first two genes in the array.

Using this approach, we successfully identify XX carriers of CVD, fully characterize a family with Bornholm eye disease, and accurately determine carrier status of an XX individual with a complex family history. Together, these results establish ONT-based targeted assembly as a clinically viable strategy for comprehensive opsin locus analysis in both XX and XY individuals, addressing a critical gap in genetic diagnostics for CVD and related conditions.

## RESULTS

### Determining *OPN1LW* and *OPN1MW* copy number using ddPCR

Probes for ddPCR were designed to target six regions in the 206 1KGP-LRSC samples analyzed in this study: *OPN1LW* exon 5, *OPN1MW* exon 5, the LCR, the promoter of the first gene in the array, the promoter of all downstream genes, and *OPN1SW* exon 1 (see Methods). Among XY samples, estimated copy numbers ranged from 0–3 for *OPN1LW* and 0–6 for *OPN1MW*. In XX samples, copy numbers ranged from 1–6 for *OPN1LW* and 1–7 for *OPN1MW* **(Table 1, Supplemental Tables 2 and 3)**.

**Table 1.** Concordance of *OPN1LW* and *OPN1MW* gene counts between ddPCR and LRS. The total number of XX samples evaluated using LRS (*n*=72) is lower than ddPCR (*n*=83) because for 9 of these samples, assembly was able to identify and resolve the LCR in only one of two X chromosome haplotypes.

| Count | XY |  |  |  | XX |  |  |  |
| --- | --- | --- | --- | --- | --- | --- | --- | --- |
|  | *OPN1LW* |  | *OPN1MW* |  | *OPN1LW* |  | *OPN1MW* |  |
|  | ddPCR | LRS | ddPCR | LRS | ddPCR | LRS | ddPCR | LRS |
| 0 | 1 | 1 | 2 | 2 | – | – | – | – |
| 1 | 117 | 117 | 53 | 56 | 2 | 2 | 2 | 2 |
| 2 | 4 | 4 | 57 | 51 | 71 | 60 | 24 | 21 |
| 3 | 1 | 1 | 7 | 9 | 7 | 8 | 31 | 27 |
| 4 | – | – | 3 | 4 | 2 | 1 | 15 | 14 |
| 5 | – | – | – | – | – | – | 7 | 4 |
| 6 | – | – | 1 | 1 | 1 | 1 | 2 | 1 |
| 7 | – | – | – | – | – | – | 2 | 1 |
| 8 | – | – | – | – | – | – | – | 2 |

### Determining *OPN1LW* and *OPN1MW* copy number using long-read sequencing (LRS)

We next evaluated the concordance of LRS with ddPCR for accurately determining the copy number of *OPN1LW* and *OPN1MW* genes. Starting with XY samples, both alignment- and assembly-based approaches were assessed to compare performance across methods.

#### Alignment-based copy number in XY individuals

For the 123 XY samples in our cohort, we first investigated the performance of alignment-based methods using two reference genomes: GRCh38, which contains 1 copy of *OPN1LW* and 3 copies of *OPN1MW*; and T2T-CHM13, which contains 1 copy of *OPN1LW* and 2 copies of *OPN1MW*.^8^ That the number of genes annotated in the reference genome is often not the same as the number of genes in any individual makes it difficult for aligners to accurately map reads to this region. Despite the longer average read lengths generated by LRS, we posited that ambiguous read mapping due to sequence similarity could hinder the ability of the alignment-based approach to accurately count gene number.

Because single nucleotide variants (SNVs) within exons 3 and 5 of the opsin genes have clinical relevance, we assessed heterozygosity at codons 171, 174, or 277 using both reference genomes **(Figure 2B)**. The presence of heterozygous SNVs in XY individuals would indicate inaccurate mapping of reads to this region. For example, if an XY individual has one copy of *OPN1LW* and *OPN1MW*, reads that span both genes and *TKTL1* (the first nonrepetitive gene downstream of the opsin array) will map to the distal end of the array. This misalignment may generate SNVs at the *OPN1MW2* position in the reference and lead to ambiguous copy number determination. Although LRS generated high-N50 reads, we still observed heterozygous SNVs with both reference genomes. Using the GRCh38 reference, we found that 1/123 XY individuals (1%) was heterozygous for an SNV in *OPN1LW*; 12/123 (10%) and 36/123 (29%) were heterozygous for a SNV in *OPN1MW* or *OPN1MW2,* respectively **(Supplemental Table 4)**. Using the T2T-CHM13 reference, no individuals were heterozygous for an SNV in *OPN1LW*, but 14/123 (11%) and 9/123 (7%) were heterozygous for a SNV in *OPN1MW* or *OPN1MW2*, respectively **(Supplemental Table 5)**.

The presence of heterozygous SNVs was most striking among the 50 XY individuals identified by ddPCR to have one copy each of *OPN1LW* and *OPN1MW*. We found that 26/50 (52%) of these individuals had at least one heterozygous SNV in *OPN1MW2* and/or *OPN1MW3* when aligned to GRCh38, and 9/50 (18%) had at least one heterozygous SNV in *OPN1MW2* when using the T2T-CHM13 reference genome. For example, ddPCR found that HG02884 carried one copy of *OPN1LW* and one copy of *OPN1MW*, while alignment identified heterozygous SNVs at codons 174 and 277 of *OPN1MW2*. Visual inspection of the alignment to GRCh38 revealed that these SNVs originated from the *OPN1LW* gene but aligned incorrectly, as several reads partially mapped to *TKTL1* **(Supplemental Figure 1)**. A reasonable, but inaccurate, interpretation of this alignment would be that the individual carries an L gene at the third position in the array and an M gene at the fourth position, resulting in an L-M-L-M orientation. Due to the difficulty of accurately mapping and quantifying L and M gene copy numbers using alignment-based approaches, we tested the accuracy of targeted *de novo* assembly for evaluating this region.

#### Assembly-based copy number in XY individuals

We used hifiasm to perform targeted assembly on all samples using reads that aligned within 1 Mb of either side of the opsin locus.^9^ We then annotated the assemblies with the LCR sequence and the *OPN1LW* and *OPN1MW* gene sequences. Using this approach across all 123 XY samples, we identified 0–4 copies of *OPN1LW* and 0–6 copies of *OPN1MW* per sample. Most individuals carried 1 copy of *OPN1LW* (117/123; 95%), while *OPN1MW* was more variable, with 56/123 (45%) having 1 copy and 51/123 (41%) having 2 copies **(Table 1)**. For one sample, HG03009, ddPCR predicted 1 L gene, while assembly generated 2 haplotypes, one that is L-M and a second that is L only. Visual inspection of these haplotypes suggested that L-M is a complete assembly while the L-only haplotype was a shorter assembly, thus we included the L-M haplotype as a correct assembly.

For *OPN1LW*, the assembly-based gene count was concordant with ddPCR for 122/123 samples (99.2%). The single discordant sample (HG00256) had 3 *OPN1LW* copies by ddPCR and 4 copies by assembly **(Supplemental Table 2)**, thus both methods indicate that the sample has more than one *OPN1LW* gene.

For *OPN1MW*, our assembly-based counts were concordant with ddPCR in 116/123 XY samples (94%). For three samples, the assemblies undercounted the number of M genes by 1; for four samples, the assemblies overcounted the number of M genes by 1 **(Table 1)**. Importantly, in no case did our assembly-based approach inaccurately count the number of L or M genes across these XY samples in such a way that would predict that an individual would be affected by CVD.

#### Assembly-based copy number in XX individuals

We then performed assembly of the 83 XX samples in the cohort. In 9 individuals, one of the two X-chromosome assemblies lacked an LCR annotation, suggesting an incomplete assembly, and these individuals were therefore excluded from further analysis. In the remaining 74 XX individuals (148 X chromosomes), we identified 1–6 copies of *OPN1LW* (summed across both haplotypes) and 0–8 copies of *OPN1MW.* 61/74 individuals (82%) had 2 copies of *OPN1LW*, while 21/74 individuals (28%) had 2 copies of *OPN1MW* and 27/74 individuals (36%) had 3 copies **(Supplemental Table 3)**. Our gene counts were concordant with ddPCR in 72/74 cases (97%) for *OPN1LW.* For the two discordant samples, the assemblies overcounted by 2 L genes in HG01893 (ddPCR predicted 2 L genes while assembly predicted 4) and undercounted by 1 in HG00236 (ddPCR predicted 4 L genes while assembly predicted 3, **Supplemental Figure 2**). For *OPN1MW*, assembly gene counts were concordant with ddPCR in 65/74 cases (87%). For the discordant counts, the assembly-based approach undercounted the number of M genes in 6/74 cases (8%) and overcounted the number of M genes in 3/74 cases (4%).

Two XX assemblies (HG01808 and HG01893) were predicted to carry only L genes and thus be affected by CVD. Because these two samples had significantly different gene counts compared to ddPCR, we suspected both haplotypes may have been incorrectly assembled **(Supplemental Table 3)**.

Assembly of HG01808 predicted both haplotypes to only have one copy of the L gene and no copies of the M genes, while ddPCR predicted 2 L and 6 M genes. To better understand this discrepancy, we manually evaluated the GRCh38 alignment results and identified six reads that partially mapped to the 5’ unique sequence of the opsin array and spanned both the annotated *OPN1LW* and *OPN1MW* genes. Among these reads, we identified multiple heterozygous SNVs in both genes, which confirmed the presence of two haplotypes with an L-M configuration in the expressed positions **(Supplemental Figure 3)**.

HG01893 was predicted by assembly to have two haplotypes with 2 L genes each and no M genes whereas ddPCR estimated 2 L and 2 M. After evaluating the alignment results, we found that no reads fully spanned the annotated *OPN1LW* and *OPN1MW* genes, but we did identify a single read spanning *OPN1MW2* and *OPN1MW3* as well as *TKTL1*. This read contained SNVs that suggested an L-M gene order, which would be expected based on the ddPCR results **(Supplemental Figure 4)**. Despite having reads spanning at least two genes in the array, both HG01808 and HG01893 were incorrectly assembled and excluded from further analysis.

### Resolving the order of the genes in the expressed position using LRS

While ddPCR can accurately count the number of opsin genes, it cannot determine their order, which is critical because gene order dictates color vision phenotype. An ideal color vision test would distinguish color-normal from color-deficient vision, identify the defect type, and measure its severity. Unfortunately, most current tests fall short in at least one of these goals. For example, an XY individual with 2 *OPN1LW* and 2 *OPN1MW* genes by ddPCR could be categorized as potentially having CVD. Resolving the order of the first two genes as L-L would confirm deutan CVD, and knowledge of the SNVs affecting the spectral-tuning amino acids in the two *OPN1LW* genes would help diagnose the severity of the CVD. Conversely, resolving the first two genes in the array as L-M would confirm the person likely has normal color vision. To determine the identity of the first two genes in the array as encoding an L or M photopigment, we annotated each haplotype with the LCR, *OPN1LW*, and *OPN1MW* reference sequences.

#### Gene order in XY individuals

In all 123 XY samples, we identified the LCR and determined the order of the first two genes in the array, with the most common order being the expected L–M orientation (119/123, 97%) **(Table 2)**. The ddPCR results indicated 8 XY samples had opsin gene counts consistent with potential inherited red-green CVD: 3 individuals lacking either an *OPN1LW* or *OPN1MW* gene (HG00556, 0L-4M; HG01170, 1L-0M; HG03788, 1L-0M) and 5 individuals with more than one L gene. Among these five, assembly showed that only one (HG00256, LLMLMLM) had L genes in the first two positions in the array, indicating true CVD **(Supplemental Figure 5)**; the other four had L-M for the first two genes in the array, consistent with normal color vision **(Table 1, Supplemental Table 6)**.

**Table 2.**
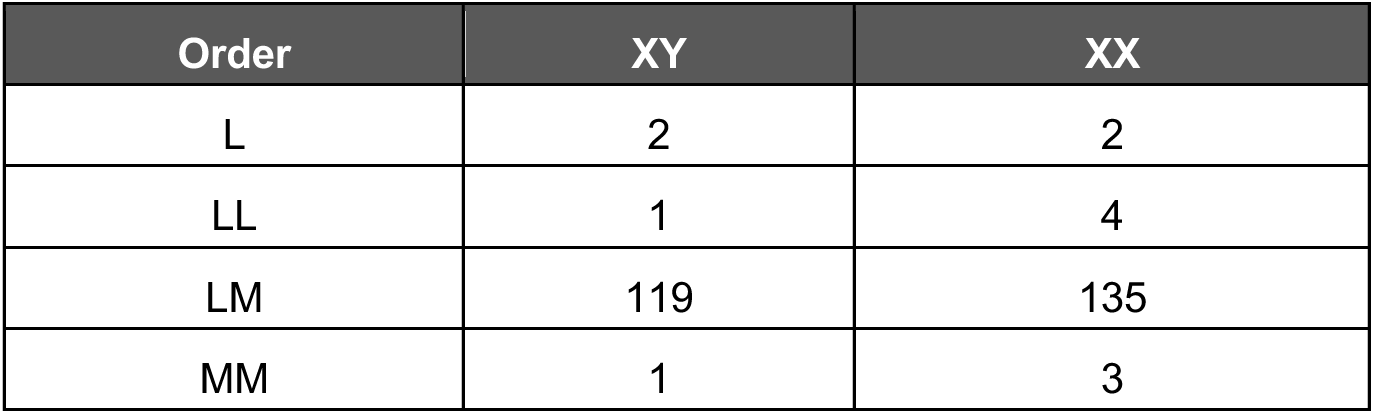
The order of the first two genes in the opsin array annotated by our assembly-based method. Data for XX samples is reported by haplotype. This excludes the 9 XX samples for which we were unable to annotate the LCR for one of the two haplotypes and 2 XX samples that we determined were incorrectly assembled.

| Order | XY | XX |
| --- | --- | --- |
| L | 2 | 2 |
| LL | 1 | 4 |
| LM | 119 | 135 |
| MM | 1 | 3 |

#### Gene order in XX individuals

Among the 72 XX individuals analyzed (144 X chromosomes), the most common order of the first two genes in the array was the L–M orientation (136/144, 94%) **(Table 2)**. However, both ddPCR and our assemblies identified four XX individuals expected to be obligate carriers of CVD, as they did not have at least 2L or 2M opsin genes. Two samples had a single L opsin gene and are thus protan CVD carriers (HG03575 and HG00231) **(Supplemental Table 7)**. Our assembly-based approach identified an M-M and an L-M-M haplotype in HG03575, while for HG00231 our assemblies identified L-M-M-M and M-M-M-M haplotypes, both of which were consistent with the ddPCR results and orthogonally validated using PCR and Sanger sequencing **(Supplemental Table 8)**.^10,11^ The other two samples (HG03808 and GM19238) had a single M opsin gene, making them obligate deutan CVD carriers. GM19238 was further found by assembly to have L genes in the expressed positions on both haplotypes, meaning this individual is predicted to be affected by deutan CVD.

In addition, ddPCR identified 9 XX individuals (GM19038, HG00097, HG00236, HG00259, HG00285, HG02555, HG02772, HG02891, HG03123) with more than 2 L genes and at least 2 M genes **(Supplemental Table 7)**, all of which could be deutan CVD carriers, depending on the positions of the L genes in the arrays. For 7 of these 9 XX samples, our assembly-based approach was concordant with ddPCR gene counts; the three discordant counts included one 4L-6M individual and two 3L-3M individuals. Assemblies for all 9 samples allowed us to resolve the order of genes on both haplotypes and determine that none were carriers of or affected by CVD.

### Concordance of assembly-based methods with optical genome mapping

A key advantage of LRS over ddPCR is the ability to determine gene order, but validating structural accuracy in complex regions is challenging because there are a limited number of orthogonal truth sets. In a prior study, PacBio HiFi sequencing of PCR amplicons was used to determine both opsin gene count and order, with results for five samples confirmed by optical genome mapping (OGM).^12^ A subsequent study used OGM to evaluate the location of a ∼697-bp segmental duplication insertion (SDIns) in the opsin locus, which was used as a primer binding site. The SDIns was evaluated across 200 alleles from both XX and XY individuals. Color vision status for this cohort was unknown, but individuals were known to carry 3 copies of the L and M opsin genes.^6^

We reasoned that if our assemblies were of high quality, we would see an SDIns distribution pattern across the locus similar to what was seen by OGM. To test this, we evaluated the position of the SDIns among 110 haplotypes from the XX and XY samples in our cohort that were predicted to have only 3 opsin gene copies **(Supplemental Table 9)**. We identified 8 different SDIns position combinations, with a pattern similar to that reported by OGM **(Supplemental Figure 6)**.^6^ For example, the previous study found the SDIns after only the third copy in the array in 76.5% of cases, and after both the first and third copy in 6% of cases. In our dataset, we observed the same pattern in 56.4% and 9% of cases, respectively, which supports the accuracy of our approach using a separate dataset.

### Concordance of targeted assembly with existing high-quality genome assemblies

We also evaluated the quality of our assemblies by comparing them to those generated by the Human Pangenome Research Consortium (HPRC) for five overlapping samples (4 XX and 1 XY).^13^ In all five cases, our assemblies were completely concordant with the HPRC assemblies **(Supplemental Table 10)**, and ddPCR counts matched both sets of assembly-based gene counts. We then used Paraphase on the HPRC PacBio HiFi data to determine gene copy number and order for each sample. We found that all were concordant with our targeted assembly and annotation approach for both copy number and the order of the first two genes in the array.

### Determining the carrier frequency of CVD using LRS

We reasoned that if our assemblies could accurately resolve the order of the first two genes in the array, we should expect to identify individuals predicted to have CVD and carriers in this cohort at frequencies consistent with published population data. Using data from a recent exhaustive meta-analysis, we estimated the prevalence of X chromosomes expected to carry a CVD-causing opsin gene array as: 4% for African ancestry (AFR), 4.5% for East Asian ancestry (EAS) and South Asian ancestry (SAS), and 8% for European ancestry (EUR). Admixed Americans (AMR) typically have a mixture of Indigenous American, AFR, and EUR ancestry with the prevalence of CVD reported as 2% in only one study; thus, we used 4% for the AMR subjects in our cohort.^3^

Our XY sample set included 29 AFR, 23 EUR, and 71 SAS, EAS, or AMR individuals, thus we expected to identify 6 XY individuals with CVD and observed 4, a frequency not statistically different from the expected prevalence (exact binomial test, two-sided *p* = 0.531). The 72 XX samples with two annotated LCRs included 17 AFR, 16 EUR, and 39 EAS, SAS and AMR individuals. Among these 72 XX individuals, we expected four carriers of a CVD-causing opsin gene array and observed six (HG00231, HG03575, HG03808, GM19238, HG00097, HG00285). This difference was not statistically significant from expected (exact binomial test, two-sided *p* = 0.296).

In total, the XY and XX samples represented 267 X chromosomes (123 from XY individuals and 144 from XX individuals), excluding nine XX samples for which one LCR could not be annotated and two XX samples with incorrect assemblies. Based on the population ancestry distribution of the cohort, we expected nine X chromosomes to harbor a genetic signature of a CVD. We observed 11 affected X chromosomes (four from XY individuals and seven affected X chromosomes from XX individuals, as sample GM19238 carried two affected X chromosomes). This difference was not statistically significant (exact binomial test, two-sided *p* = 0.52).

### Long-read sequencing and targeted assembly resolves a case of Bornholm eye disease

Based on the success of targeted assembly for accurately identifying X chromosomes with CVD in XX and XY samples, we applied the method to a challenging clinical case. We identified a family in which three of five XY siblings had been previously diagnosed with Bornholm eye disease, a rare X-linked condition characterized by red-green CVD and high myopia.^14–16^ ^17,18^ Adaptive optics retinal imaging had shown an abnormal cone photoreceptor phenotype, and previous studies found that while unaffected XY siblings carried 1 L and 2 M genes, the affected XY siblings carried 2 L and 2 M opsin genes. However, the order of these genes within the array could not be resolved. For one of the siblings, amplification and sequencing of the first gene in the array (the LCR-proximal gene) using previously described methods had shown that it encoded an L cone opsin.^11^ Exon 3 of this gene was known to have a rare combination of SNVs (denoted LVAVA for the amino acids encoded at the variant positions) that causes exon 3 skipping during mRNA processing and is associated with high myopia. Because both L genes had been amplified and sequenced together, their sequences were determined by deduction, but the location of the second L gene within the array remained unknown.

To clarify the position and sequence of all genes, we performed targeted LRS on blood-derived DNA from the proband and one unaffected XY sibling. Assembly of the proband revealed an L-L-M-M gene order, while the unaffected sibling carried an L-M-M array. An XY individual with an L-L-M-M array will only express L and S opsin, but not M opsin, and thus will have CVD. Importantly, we confirmed that exon 3 of the first L gene in the array of the proband did encode the rare LVAVA variant combination that causes exon 3 skipping, while exon 3 of the second L gene encodes the MVVVA variant, which has also been associated with high myopia **(Figure 3)**.^4,19^ Thus, using LRS and targeted assembly, we were able to molecularly explain both the CVD and myopia phenotypes.

**Figure 3.**
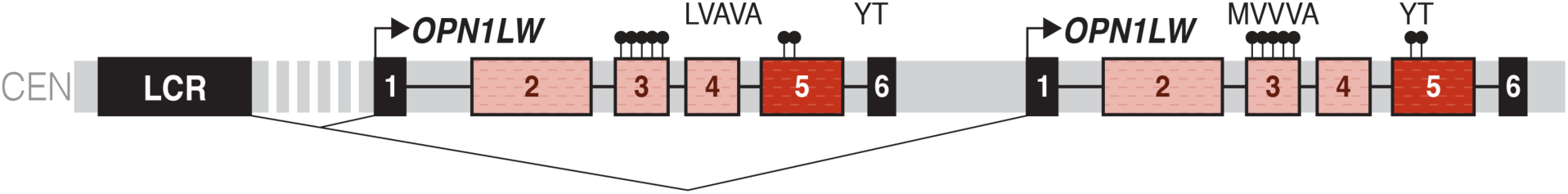
Clinical application of targeted long-read sequencing of the opsin locus. Targeted long-read sequencing and assembly of the opsin locus of the proband from a family with Bornholm eye disease revealed an L–L–M–M gene order. The first expressed gene carries the LVAVA exon 3 haplotype that causes exon 3 skipping, while the second gene carries the MVVVA haplotype. Both genes encode L-class photopigments and harbor splice-disrupting variants, explaining the combination of color vision deficiency and high myopia observed in affected individuals from the family.

### Resolving gene order and count in an individual suspected to be a double carrier of CVD

An XX individual who carries a protan array on one X chromosome and a deutan array on the other will have normal color vision, but all of their XY offspring will be affected by red-green CVD.^20^ We identified an XX individual suspected to be a double carrier based on family history. Her father and brother both had CVD, making her mother an obligate carrier. Additionally, her daughter was known to carry only one L gene by ddPCR, establishing the daughter as an obligate protan carrier. This pedigree structure indicates that the proband inherited an affected X chromosome from her father and was at risk for inheriting an affected X chromosome from her mother as well. We hypothesized that the proband carried one X chromosome with a deutan signature (L-L or L) and one with a protan signature (M-M or M)—an arrangement that would result in expression of both L and M opsin pigments and confer normal trichromatic color vision despite carrying two CVD-causing haplotypes.

We performed targeted LRS and assembly on this individual and identified two affected haplotypes: one with an L-L-M gene order and another with an M-M gene order, confirming our hypothesis. Notably, ddPCR alone would have identified this individual as having 2L and 3M genes, which, given her normal color vision, would have incorrectly designated her as a non-CVD carrier. This again illustrates the unique capability of our assembly-based method to determine carrier status by resolving both gene count and order in complex scenarios where conventional methods would yield misleading results.

## DISCUSSION

We demonstrate that LRS combined with targeted *de novo* assembly provides a comprehensive, reference-free method for evaluating the human opsin gene cluster. Across 195 individuals from diverse populations, our assembly-based approach achieved high concordance with ddPCR for gene copy number in XY individuals (99% for *OPN1LW* and 94% for *OPN1MW*), with slightly lower rates in XX individuals (97% and 88%, respectively). Our method also resolves gene order, an essential detail that is currently inaccessible to existing molecular assays. This enabled us to more accurately predict color vision phenotype. The frequency of CVD-associated genotypes in our cohort—3.3% in XY individuals and 6.3% in XX individuals—matched population-based estimates from color vision testing, supporting both the accuracy of our approach and its potential as a clinical diagnostic test.

Our assembly-based approach was essential as reference-guided evaluation can result in incorrect or misleading alignments. Despite an average read N50 of 33.7 kbp **(Supplemental Table 1)**, alignment to either GRCh38 or T2T-CHM13 produced spurious heterozygous variants in hemizygous XY individuals at clinically important codons. These artifacts can arise because aligners anchor reads using flanking unique sequences, forcing internal opsin gene sequences to map to whichever copy is present in the reference regardless of the individual’s true gene content. Visual inspection of these alignments shows how reads spanning the terminal unique gene, *TKTL1*, were assigned to incorrect opsin copies, creating false heterozygosity that could lead to erroneous interpretation of gene count or order. These findings demonstrate that accurate interrogation of the opsin locus requires reference-free reconstruction and would argue that improvements in alignment algorithms or reference genome completeness cannot resolve the fundamental problem of mapping reads to a locus whose structure varies substantially among individuals.

The clinical utility of our approach extends beyond copy number to several capabilities not achievable with existing methods. First, by resolving the identity of genes in the two expressed positions, we can distinguish individuals whose copy number suggests potential CVD from those whose gene arrangement confers normal color vision. In our cohort, five XY individuals had more than one L gene by ddPCR, but only one had L genes in both expressed positions and would therefore have CVD, a distinction impossible without order information. Second, this method enables carrier detection in XX individuals, who have been largely excluded from molecular diagnosis because it is impossible to distinguish the two X-chromosome arrays by PCR or other quantitative methods. We identified six carriers of CVD among 72 fully resolved XX individuals, including one who carried CVD-causing arrangements on both X chromosomes and is therefore affected by CVD despite having six L genes and one M gene by ddPCR. Third, the ability to extract complete coding sequences from assembled genes allows identification of pathogenic variants and prediction of phenotypic severity, as demonstrated in the Bornholm eye disease family where we confirmed the presence of exon 3 splice-affecting variants in both expressed L genes.

The Bornholm eye disease family illustrates the value of this approach for clarifying the molecular mechanisms underlying clinical phenotypes. Previous studies had established that affected individuals carried two L genes and that the first encoded the LVAVA haplotype associated with exon 3 skipping and high myopia, but the position and sequence of the second L gene remained unknown. Our assembly showed that the second expressed gene also carries an L-class splice-disrupting haplotype (MVVVA).^15,16^ The presence of two L-class genes with compromised expression provides a mechanistic explanation for the CVD, unusually severe myopia, and abnormal cone photoreceptor phenotype observed in this family.

Our ability to resolve both X-chromosome arrays in XX individuals further demonstrates the unique diagnostic value of our approach. In a suspected double carrier with normal color vision, ddPCR results found 2L and 3M genes, which could be interpreted as indicating non-carrier or single-carrier status depending on family history. Our assembly revealed she carries an L-L-M array on one X chromosome and an M-M array on the other, a combination that produces normal trichromatic vision but guarantees that all XY offspring will have CVD. This result has direct implications for genetic counseling and reproductive planning.

Our study has several limitations. First, assembly quality can depend on multiple factors. Nine XX samples (10.8%) could not be fully resolved because one haplotype lacked an annotated LCR, and two additional samples produced assemblies discordant with ddPCR despite having reads spanning multiple genes. These failures may reflect stochastic under-sampling of one haplotype or insufficient read lengths for accurate phasing and may be addressable through higher coverage, adaptive sampling enrichment, or improved assembly algorithms. Although, it is important to note that we were able to generate complete assemblies for samples with lower coverage, read N50 values, and average read lengths than some of the 11 excluded samples **(Supplemental Figure 2B)**. Second, concordance for *OPN1MW* copy number was lower than for *OPN1LW*, particularly in XX individuals, suggesting that arrays containing multiple M genes are more susceptible to assembly fragmentation. This may reflect the paucity of distinguishing sequence features among M gene copies compared with those distinguishing L and M genes. Finally, although ONT accuracy has improved substantially, systematic errors in homopolymer regions remain a consideration when interpreting SNVs, and orthogonal confirmation may be warranted for novel variants in clinical samples.

The use of samples from the 1000 Genomes Project also introduces several caveats.^21,22^ First, individuals were self-reported to be healthy. Individuals known to have CVD or a family history of CVD may have self-excluded from the study, potentially lowering the observed frequency of CVD haplotypes in our dataset. Second, we were unable to perform behavioral confirmation of CVD or review family history among those individuals for whom sequencing predicts the presence of a CVD haplotype. Finally, DNA used for sequencing of these samples was derived from lymphoblastoid cell lines, so we cannot exclude the possibility that some of the CVD-associated haplotypes arose from replication errors during cell propagation.

Further optimization could enhance clinical implementation of this approach. Adaptive sampling strategies that enrich for the opsin locus during sequencing could improve assembly completeness and reduce the number of haplotypes that were inaccurately or not completely assembled.^23^ Integration of our approach with automated variant annotation pipelines could facilitate identification and interpretation of coding variants in *OPN1LW* and *OPN1MW*. Extension of this framework to other structurally complex, clinically relevant loci such as *SMN1*/*SMN2*, *CYP2D6*, and the immunoglobulin and HLA regions is a possible future application of the methods developed here.^24^

In conclusion, we have shown that ONT-based targeted assembly provides a robust, scalable method for comprehensive analysis of the human opsin gene cluster in both XY and XX individuals. By enabling simultaneous determination of gene number, order, and coding sequence on both X chromosomes, this approach addresses a longstanding diagnostic gap for inherited CVD and X-linked cone dysfunction syndromes. As LRS becomes increasingly accessible in clinical laboratories, assembly-based characterization of paralogous gene families is poised to become an essential component of clinical genetic testing.

## METHODS

### Sample selection

Publicly available data from 206 of the 1000 Genomes Project samples generated by the 1KGP-LRSC and sequenced on the R10.4.1 pore were used for analysis.^7,21,22^ These samples had an average genome-wide depth of coverage of 37x, and average read N50 of 33.7 kbp **(Supplemental Figure 2)**. Selected samples included individuals from all five 1KGP superpopulations, which included 83 XX samples (18 AFR, 12 AMR, 18 EAS, 18 EUR, 17 SAS) and 123 XY samples (29 AFR, 19 AMR, 17 EAS, 23 EUR, 35 SAS).

### Ethics declaration

Non-1KGP samples sequenced as part of this study were deidentified and determined by the University of Washington Institutional Review Board not to constitute human subjects research (exemption #35386).

### Droplet digital PCR

Droplet digital PCR was performed on all 1KGP samples used in this study. We used 8 primers and 6 probes to analyze the opsin gene array structure for all samples **(Supplemental Table 11)**. This allowed us to distinguish between the first and downstream genes in the array using probes targeting the promoter region, which differs between these positions. While this region cannot differentiate between *OPN1LW* and *OPN1MW* genes, it identifies positional information within the array. Each array typically contains one first gene; however, rare mutations affecting the distinguishing SNP can result in “0 first gene” reports. To distinguish between L and M opsin genes, primers 1 and 2 along with FAM and HEX probes target a SNP at codon 309 in exon 5. This codon was selected because it is closely linked to codons 277 and 285, which control approximately 21 nm of the spectral sensitivity difference between L and M cone photopigments; yet unlike these positions, codon 309 lacks surrounding silent polymorphisms that complicate ddPCR assays. Previous studies found mutations at position 309 in only 5 of 869 normal trichromatic XY individuals.^11^

Up to 300 ng of genomic DNA was used in each 22-μL reaction, which contained ddPCR Multiplex Supermix (Bio-Rad), primers at 950 nM final concentration, and probes at 250 nM final concentration. Primers 1–6 and the FAM, HEX, Cy5, and Cy5.5 probes were synthesized by IDT, while primers 7–8 and the ATTO590 probe were obtained from Bio-Rad. Reactions also included 2–5 units each of BamHI and EcoRI restriction enzymes (NEB) to separate *OPN1LW* and *OPN1MW* genes onto different DNA fragments, ensuring each droplet theoretically contained 0 or 1 target molecule.

Droplets were generated using the AutoDG system (Bio-Rad) with droplet generation oil following manufacturer’s instructions, producing a final PCR volume of 40 μL. Thermal cycling was performed immediately using a C1000 deep-well thermal cycler (Bio-Rad) with the following conditions: initial denaturation at 95°C for 10 minutes, followed by 40 cycles of 94°C for 30 seconds and 62.2°C for 1 minute with a 2°C/second ramp rate, and final denaturation at 98°C for 10 minutes before cooling to 4°C. Droplets were analyzed immediately after PCR using the QX600 Droplet Reader (Bio-Rad) in direct quantitation mode across six channels: FAM for *OPN1MW* exon 5, HEX for *OPN1LW* exon 5, Cy5 for the first gene promoter, Cy5.5 for downstream gene promoters, ROX for the LCR, and ATTO590 for OPN1SW exon 1. All probes used BHQ-1 as quencher except ATTO590, which used Iowa Black.

For data analysis, the QX600 software calculated DNA concentration in copies per μL for each probe. Gene copy numbers were determined by normalizing to the LCR, a unique 36-bp element that exists as a single copy per X-chromosome opsin array.^25^ The *OPN1SW*/LCR ratio is expected to be approximately 2 for XY individuals, who have two autosomal *OPN1SW* genes but only one X chromosome, and approximately 1 for XX individuals, who have two of each. The relative ratios of opsin-specific probes to LCR provided the number of *OPN1LW* and *OPN1MW* genes after correcting for X-chromosome number.

### Targeted long-read sequencing

We performed targeted LRS using adaptive sampling for the two siblings from the family affected by Bornholm eye disease and for the XX individual presumed to be a double carrier of CVD. For all three individuals, high molecular weight DNA was extracted from whole blood using a Qiasymphony. DNA was quantified using a Qubit Fluorometer and fragment size was evaluated using an Agilent FEMTO Pulse. Libraries for sequencing were prepared using the SQK-LSK114 library kit (ONT). Samples were sequenced on the PromethION platform using the R10.4.1 pore. Adaptive sampling was performed using the T2T-CHM13 reference genome with target regions specific to the opsin cluster (chrX:151317896-153570523 [*OPN1LW*/*OPN1MW*]), the autosomal *OPN1SW* gene (chr7:129984638-130187947 [*OPN1SW*]), and 3 control genes (chr15:21319888-21521793 [*NDN*], chr17:50951162-51168680 [*COL1A1*], chrX:146076547-146315797 [*FMR1*]). Each sample was sequenced for approximately 24 hours before washing and loading of a second sample.

### Alignment and variant calling

Reads were aligned to both the GRCh38 and T2T-CHM13 reference genomes using minimap2 (v2.28) with the map-ont preset.^26^ Variant calling and phasing was performed using Clair3 (v1.0.8) with the ONT R10.4.1 model and --enable_phasing option.^27^ Aligned BAM files were haplotagged using Longphase (v1.7.3).^28^ BAM file manipulation was performed using Samtools (v1.19).^29^ To assess alignment accuracy, a custom Python script was used to extract genotypes at clinically relevant codons (153, 171, 174, 178, 180, 277, and 285) within the opsin genes and identify samples with heterozygous SNV calls, which indicate mapping ambiguity in hemizygous XY individuals.

### Assembly and annotation

Targeted haplotype-resolved *de novo* genome assemblies were generated using reads that mapped to a 2-Mb region around the opsin array. Coordinates used were chrX:153121316-155216212 (GRCh38). Reads were extracted from minimap2-aligned BAM files using Samtools view with the -F 0x900 flag to retain only primary alignments.^29^ Assemblies were generated using hifiasm (v0.25.0-r726) with the ’--ont’ option for ONT data and the ’--hg-size’ flag set to 2m (megabases).^9^

Annotation of assembly files was performed using Exonerate (v2.4.0) with the protein2genome model.^30^ Reference sequences used for annotation included: (1) a 1.5-kb fragment containing the LCR (GenBank accession NC_000023.11, chrX:154,139,711-154,141,275); (2) the *OPN1LW* exon 5 reference sequence (NM_020061.6, chrX:154,156,293-154,156,534); and (3) the *OPN1MW* exon 5 reference sequence (NM_000513.2, chrX:154,193,408-154,193,647). Exonerate was run with parameters --bestn 10 --showtargetgff TRUE to identify all instances of each reference sequence within each assembled haplotype.

### Gene classification

Assembled opsin genes were classified as *OPN1LW* (L) or *OPN1MW* (M) based on the amino acid encoded at position 277 in exon 5, which is a primary determinant of spectral tuning. Genes encoding tyrosine at position 277 were classified as L genes, while genes encoding phenylalanine were classified as M genes. This classification was performed by extracting the codon 277 sequence from each annotated gene in the assembly and translating to the corresponding amino acid. In cases where exon 5 sequence quality was insufficient for confident base calling, genes were classified based on the amino acid at position 285 (alanine for L genes, threonine for M genes) or by overall sequence similarity to the *OPN1LW* and *OPN1MW* reference sequences across the full coding region.

### Gene order determination

The order of opsin genes within each assembled haplotype was determined by annotating the position of the LCR and each opsin gene, then ordering the genes by their genomic coordinates relative to the LCR. For each haplotype, the first gene (Gene 1) was defined as the opsin gene nearest the LCR on its 3’ side, with subsequent genes numbered sequentially in the direction away from the LCR. The identity of genes in the first two positions (the “expressed positions”) was recorded for each haplotype, as only these genes contribute to color vision phenotype. Arrays were classified as consistent with normal color vision if the first two positions contained one L gene and one M gene (L-M order), and as consistent with CVD if both expressed positions contained genes of the same class (L-L for deutan CVD, M-M for protan CVD).

### Assembly quality assessment

Assemblies were assessed for completeness by verifying the presence of annotated features including the LCR and at least one opsin gene per haplotype. For XX samples, both haplotypes were required to have an annotated LCR for inclusion in downstream analyses; samples with only one annotated LCR (*n*=9; HG00154, HG00250, HG00513, HG01113, HG01841, HG01973, HG02837, HG03752, HG04202) were excluded from gene order and carrier status analyses. Assemblies were flagged as potentially erroneous when the total gene count differed from the ddPCR estimate by more than one gene. For flagged assemblies, reads were manually inspected in IGV (Integrative Genomics Viewer) to assess whether the assembly accurately represented the underlying read data.^31^ Assemblies were classified as incorrect if manual inspection revealed that reads spanning multiple genes supported a gene count or order inconsistent with the assembly output. Two XX samples (HG01808 and HG01893) were identified as misassembled based on this manual review and were excluded from carrier frequency analyses.

### Determining the position of SDIns within the opsin array

To validate assembly accuracy, we analyzed the distribution of a ∼697-bp SDIns previously characterized by optical genome mapping.^6,12^ The SDIns sequence was identified in assemblies by BLAST alignment of the SDIns reference sequence (chrX:154,239,698-154,240,398, hg38) against each assembled haplotype using blastn with an e-value threshold of 1e-10 and requiring ≥95% sequence identity over ≥90% of the query length. The position of each SDIns relative to the opsin genes in the array was recorded (e.g., after Gene 1, after Gene 2, after Gene 3). The distribution of SDIns positions was compared to previously published OGM data from 200 haplotypes carrying 3 opsin genes. Analysis was restricted to samples in our cohort with 3 total opsin genes as determined by ddPCR (*n*=110 haplotypes).

### Paraphase analysis

To further validate our assembly-based approach, we compared our results to Paraphase analysis of PacBio HiFi data for five samples that overlap between our cohort and the HPRC.^13^ Paraphase (v3.1.1) was run on HPRC PacBio HiFi BAM files using default parameters.^5^ Paraphase output for the opsin locus was compared to our ONT-based targeted assembly results for concordance of gene copy number and the identity of genes in the first two positions of the array.

### Comparison with HPRC assemblies

For five samples present in both our cohort and the HPRC dataset, we compared our targeted assemblies to the high-quality phased genome assemblies generated by the HPRC (hprc_r2_v1.0) using PacBio HiFi and ONT data.^13^ HPRC assemblies were downloaded from https://data.humanpangenome.org/. The opsin locus was extracted from each HPRC haplotype assembly and annotated using the same Exonerate-based pipeline applied to our targeted assemblies. Gene count and order were compared between HPRC assemblies, our targeted ONT assemblies, and ddPCR results.

### Statistical analysis

Concordance between ddPCR and assembly-based gene counts was calculated as the percentage of samples for which the two methods produced identical counts for *OPN1LW* and *OPN1MW*, evaluated separately for XY and XX individuals. To assess whether the observed frequency of CVD-associated genotypes in our cohort was consistent with population-based estimates, we used two-sided exact binomial tests implemented in Python. Expected frequencies were calculated using published population-specific CVD prevalence estimates.^4,19^ For XX individuals, the expected carrier frequency was set equal to the population-specific CVD prevalence, as carrier frequency approximates affected frequency when the trait is X-linked and relatively common. Expected counts were calculated by summing the product of sample size and expected frequency for each population. *P*-values less than 0.05 were considered statistically significant.

## DATA AVAILABILITY

LRS data for 1KGP samples is available at https://s3.amazonaws.com/1000g-ont/index.html. Data for individuals sequenced as part of this study are available from the corresponding author.

## CODE AVAILABILITY

Code used in this study can be found on GitHub at: https://github.com/millerlaboratory/OPSIN-ASSEMBLY-ANNOTATION

## ACKNOWLEDGEMENTS

DEM is supported by the National Institutes of Health through the NIH Director’s Early Independence Award DP5OD033357.

## AUTHOR CONTRIBUTIONS

ZBA: conceptualization, formal analysis, investigation, software, writing

TP: formal analysis, software

ND: formal analysis, software

SHRS: investigation

JG: investigation

ALM: writing

JC: resources, writing

MN: conceptualization, resources, formal analysis, supervision, writing

DEM: conceptualization, resources, formal analysis, supervision, writing

