## Supplemental Figures for "Long-read sequencing with targeted assembly of the opsin locus accurately evaluates genes in expressed positions"

### Supplementary Figures

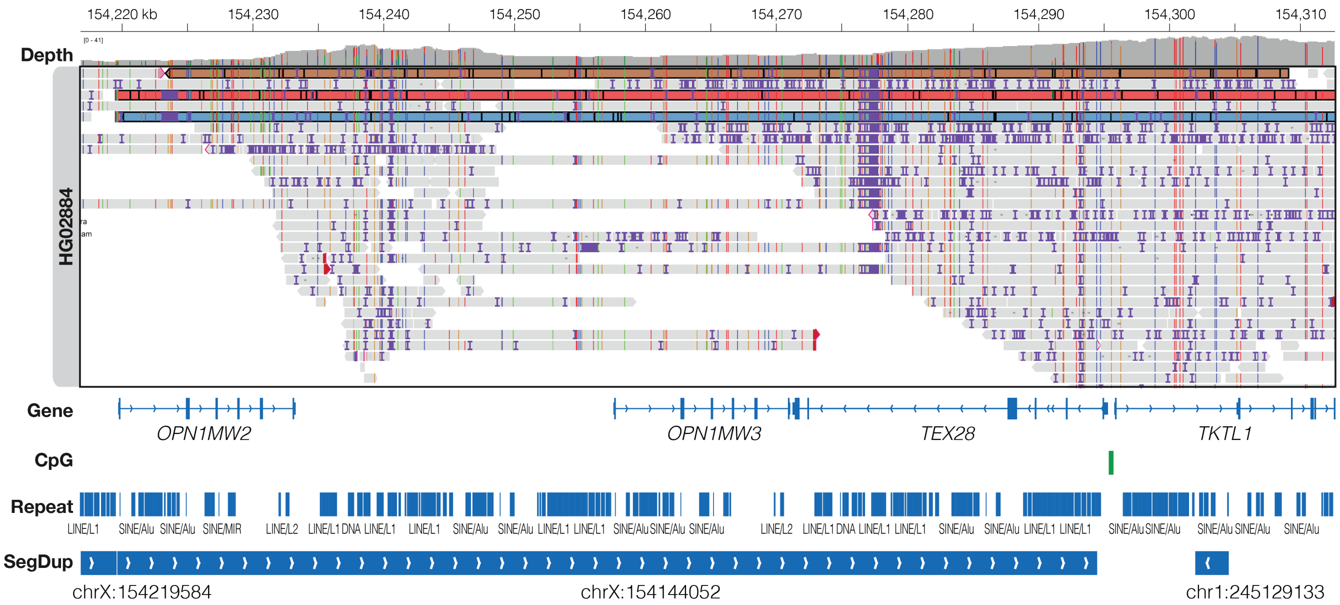


**Figure S1. Misalignment of opsin genes in HG02884.** IGV view of long-read sequencing (LRS) data aligned to the GRCh38 reference genome. The highlighted reads contain heterozygous single nucleotide variants (SNVs) assigned to *OPN1MW2*, but these reads are actually derived from *OPN1LW* and *OPN1MW*. Because HG02884 carries only two opsin genes—compared with four in the reference—reads that extend into the first nonrepetitive downstream gene (*TKTL1*) anchor there and are consequently misaligned more distally within the array.

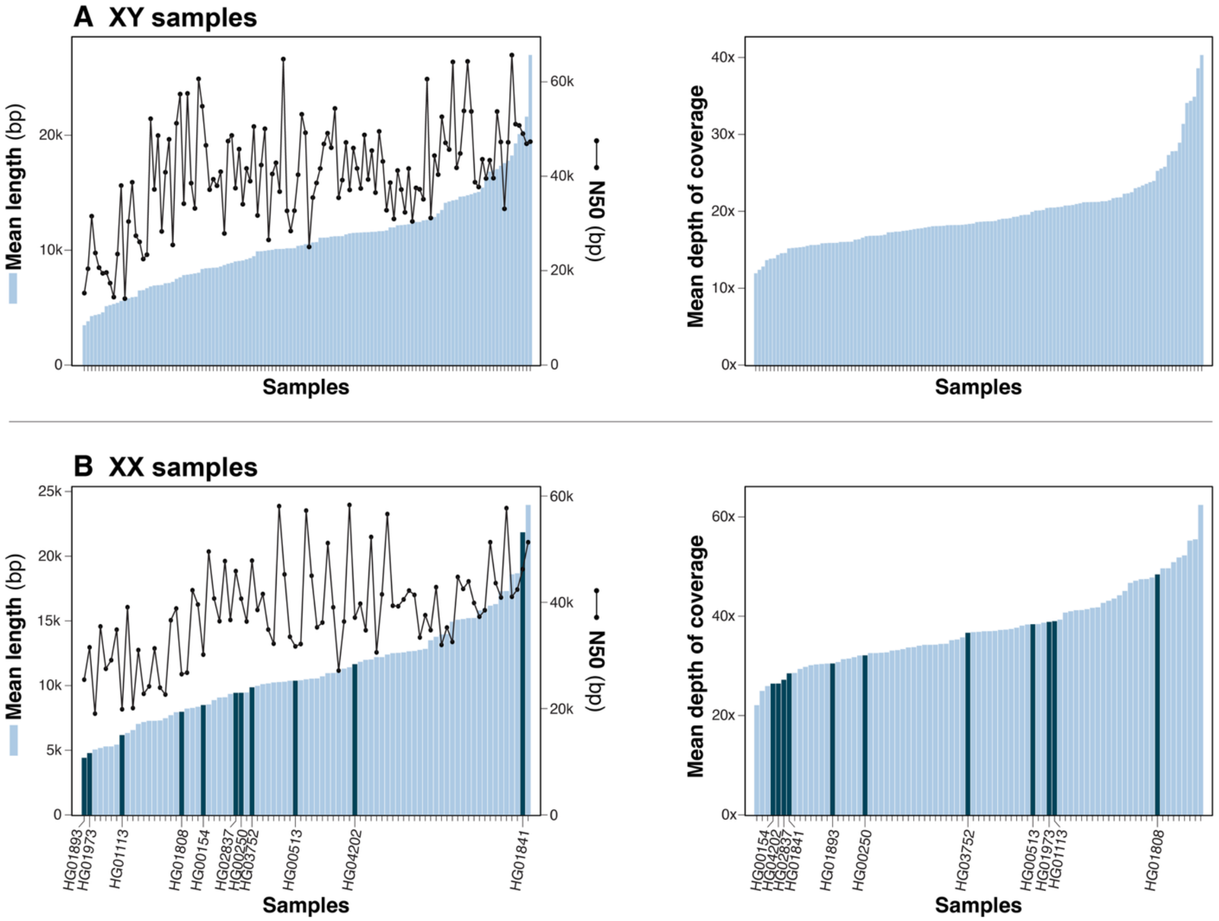


**Figure S2. Per-sample X-chromosome sequencing metrics in 206 samples.** Mean read length, read N50, and mean depth of coverage were generated using cramino and samtools for the 123 XY samples **(A)** and 83 XX samples **(B)** evaluated in this study. 11 XX samples (dark blue) were excluded from analysis due to incorrect or incomplete assembly of one X chromosome. Nine of these (HG00154, HG00250, HG00513, HG01113, HG01841, HG01973, HG02837, HG03752, HG04202) lacked assembly of one of their LCRs, while two of them (HG01893 and HG00236) had discordant *OPN1LW* gene counts between ddPCR and LRS assembly.

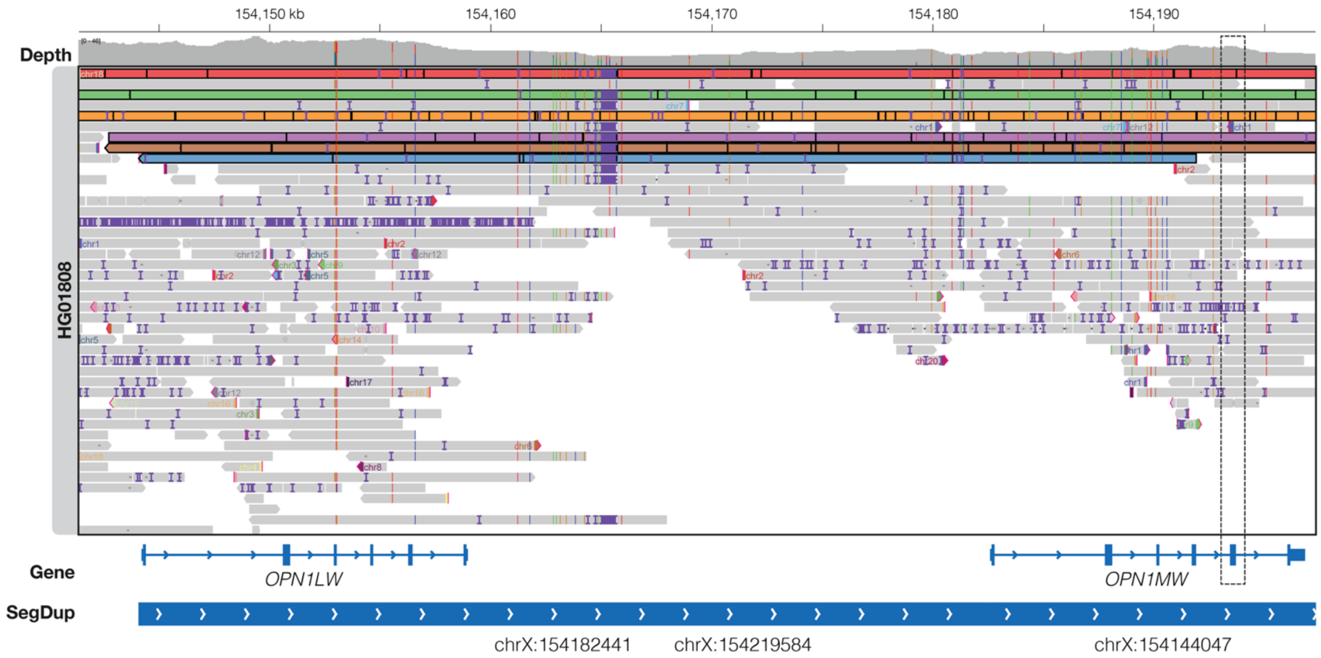


**Figure S3. Aligned reads support ddPCR copy number estimate and confirm an assembly error in HG01808.** An IGV view of ONT reads aligned to the GRCh38 reference genome. ddPCR estimated that this XX sample carried two *OPN1LW* genes and six *OPN1MW* genes, whereas LRS assembly and annotation estimated four total L genes (two per haplotype) and no M genes. When aligned to GRCh38, the highlighted reads begin outside the repetitive region and span the first two genes in the array. Compared to the reference, exon 5 of the second opsin gene in these reads (dashed box) lacks L-specific SNVs, indicating that this copy corresponds to an M gene. This supports the presence of two L and at least two M genes, consistent with the ddPCR estimate.

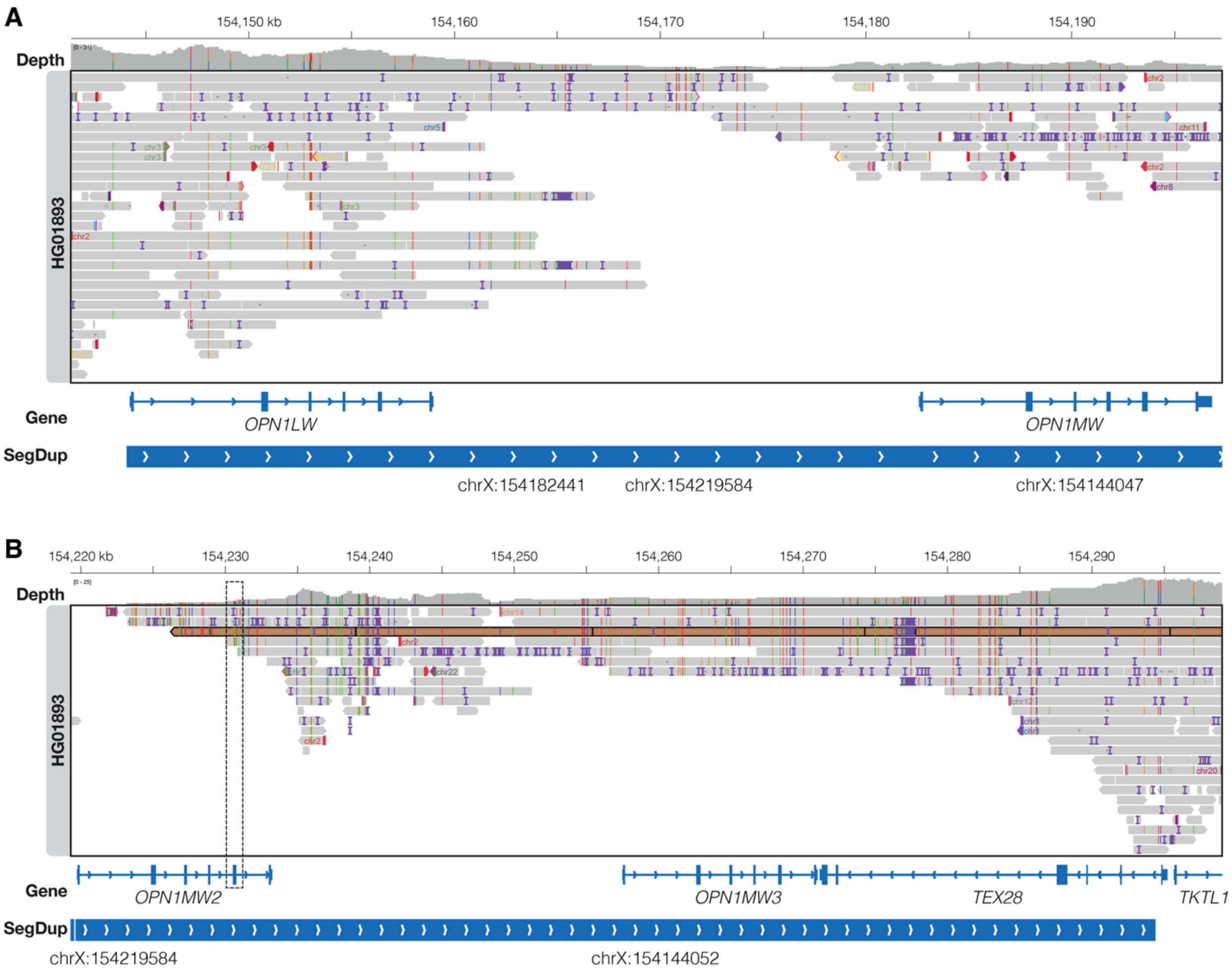


**Figure S4. A distal opsin array read supports the gene structure inferred from assembly and annotation for HG01893.** IGV views of ONT data aligned to the GRCh38 reference genome. Although no reads spanned the first two genes in the opsin array **(A)**, the read highlighted in orange **(B)** spans the last two opsin genes (*OPN1MW2* and *OPN1MW3*) as well as *TEX28* and *TKTL1* (the first gene outside the repetitive region). The aligner used the portion of the read mapping to *TKTL1* to anchor the read there and tolerates the resulting SNVs in the opsin genes. However, SNVs in exon 5 of *OPN1MW2* (dashed box) indicate that it is an L gene, while SNVs in exon 5 of *OPN1MW3* suggest it is an M gene.

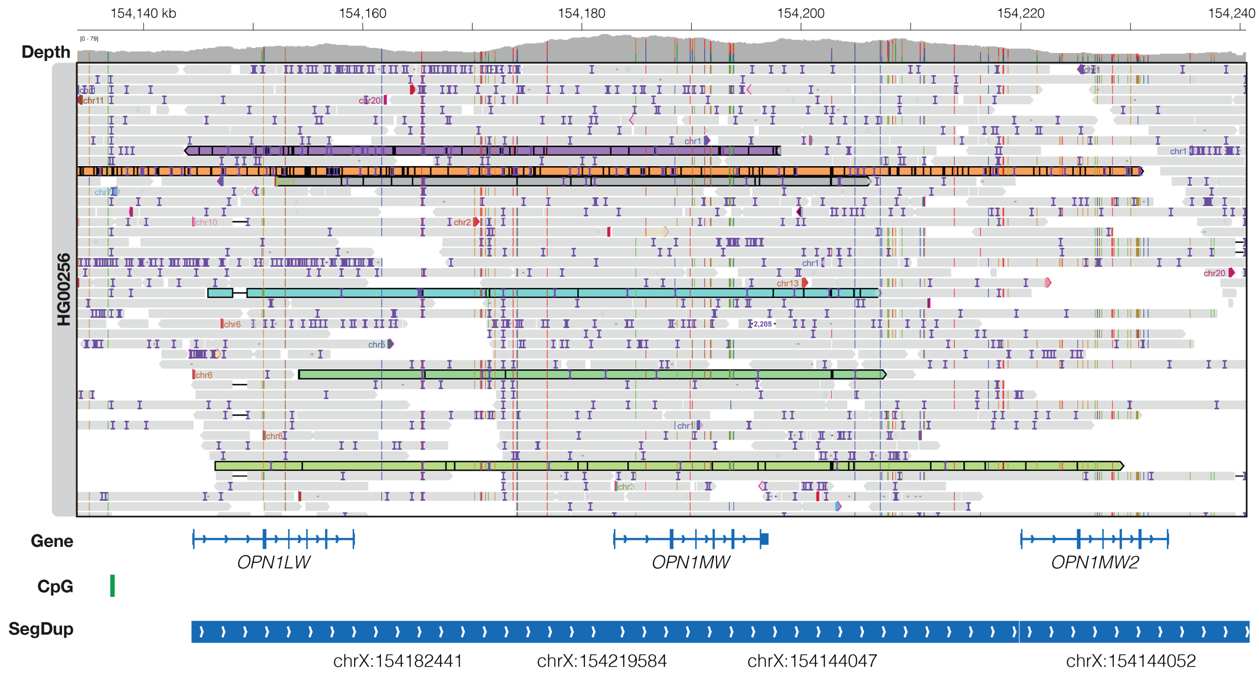


**Figure S5. XY sample with suspected CVD has aligned reads that support the assembled gene order.** An IGV view of LRS data aligned to GRCh38. The highlighted reads overlap two or more genes at Xq28 and contain SNVs in exon 5 of *OPN1MW* that support a gene order of L-L in the expressed positions.

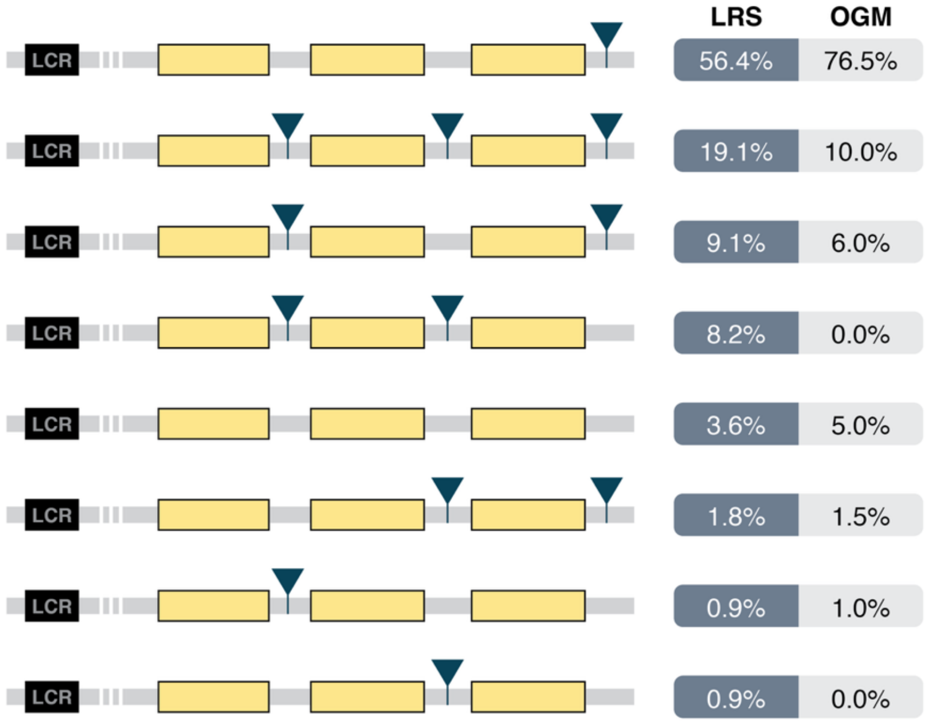


**Figure S6. Distribution of the ~697-bp segmental duplication insertion (SDIns) within the opsin gene array.** Assembled haplotypes from individuals predicted to carry three total opsin genes were analyzed to determine the position(s) of the previously described SDIns relative to the opsin genes. In each assembly, SDIns positions (blue triangles) were identified by sequence alignment and annotated relative to gene order within the array (e.g., after Gene 1, Gene 2, or Gene 3). The frequency of each SDIns positional configuration observed in our LRS dataset is shown. This distribution closely mirrors prior findings obtained by optical genome mapping (OGM) (Haer-Wigman *et al*., 2024), supporting the structural accuracy of the targeted assembly approach for resolving the opsin locus.
